# Outbreak or pseudo-outbreak? Integrating SARS-CoV-2 sequencing to validate infection control practices in an end stage renal disease facility

**DOI:** 10.1101/2020.12.30.20249062

**Authors:** Bridget L. Pfaff, Craig S. Richmond, Arick P. Sabin, Deena M. Athas, Jessica C. Adams, Megan E. Meller, Kumari Usha, Sarah A. Schmitz, Brian J. Simmons, Andrew J. Borgert, Paraic A. Kenny

## Abstract

**Background:** The COVID-19 pandemic of 2020 poses a particularly high risk for End Stage Renal Disease (ESRD) patients and led to a need for facility-wide control plans to prevent introduction and spread of infection within ESRD facilities. Rapid identification of clusters of contemporaneous cases is essential, as these may be indicative of within-facility spread. Nevertheless, in a setting of high community COVID-19 prevalence, a series of ESRD patients may test positive at around the same time without their shared ESRD facility being the nexus for disease spread. Here we describe a series of five cases occurring within an eleven-day period in November 2020 in a hospital-based 32-station ESRD facility in southwest Wisconsin, the subsequent facility-wide testing, and the use of genetic sequence analysis of positive specimens to evaluate whether these cases were linked.

**Methods:** Four patient cases and one staff case were identified in symptomatic individuals by RT-PCR. Facility-wide screening was initiated at the request of local public health and conducted using Abbot BinaxNOW antigen tests. SARS-CoV-2 genome sequences were obtained from residual diagnostic test specimens using an amplicon-based approach on an Ion Torrent S5 sequencer.

**Results:** Residual specimens from 4 of 5 cases were available for sequence analysis. Each sequence was very clearly genetically distinct from the others, indicating that these contemporaneous cases were not linked. Facility-wide screening of 47 staff and 107 patients did not identify any additional cases.

**Conclusions:** These data indicate that despite the outward appearance of a case cluster, the facility did not experience within-facility spread nor serve as the epicenter of a new outbreak, suggesting that the enacted rigorous infection control procedures (screening, masking, distancing) practiced stringently by patients and staff were sufficient to permit dialysis to proceed safely in a very high-risk population under pressure from increasing community spread. These data also demonstrate the utility of rapid turnaround SARS-CoV-2 sequencing in outbreak investigations in settings like ESRD facilities.

## INTRODUCTION

A global pandemic of novel coronavirus, SARS-CoV-2, was declared by the World Health Organization on March 11, 2020. The first reported death from COVID-19 in the United States was an End Stage Renal Disease (ESRD) patient (1). Accumulating data show that ESRD patients are at higher risk of adverse outcomes when infected with the virus (2, 3); however, they still depend on regularly scheduled treatments to maintain their health. Detailed guidance on optimal control measures to contain COVID-19 in dialysis is available and emphasizes staff and patient education, early screening of patients, managing patients with symptoms or illness, managing resources and managing the workforce (4-6). There are limited protocols and procedures in place to guide facility-wide testing efforts, and some suggest transferring these patients to designated COVID-19 facilities or hospitals in response to identification of cases (4).

Because of the risk of COVID-19 to ESRD patients and the risk of subsequent spread to other vulnerable populations (7), rapid detection and prevention of COVID-19 spread within dialysis facilities is of critical importance. In Wisconsin, two cases occurring within seven days in an ESRD facility is considered an outbreak warranting public health investigation (8). As cases in the community become more widespread, the probability of two or more unrelated cases utilizing the same ESRD facility increases. Efficiently distinguishing such “pseudo-outbreaks” from true cases of intra-facility spread may allow more efficient use of both infection control and public health staff resources, as well as providing reassurance to both staff and patients about the actual effectiveness of infection control measures employed.

Here we describe a series of five cases occurring within an eleven-day period in a hospital-based 32-station ESRD facility in southwest Wisconsin, the subsequent facility-wide testing, and the use of genetic analysis of positive specimens to unambiguously demonstrate that these cases were not linked to spread of a commonly circulated virus within the facility.

## MATERIALS AND METHODS

### SARS-CoV-2 testing

Each of the five cases defining the potential cluster was diagnosed by RT-PCR from nasopharyngeal specimens at Gundersen Health System laboratories. Facility-wide surveillance testing was performed using anterior nares swabs with the Abbott BinaxNOW antigen test kit. *SARS-CoV-2 sequencing and analysis:* cDNA was generated from residual RNA from diagnostic specimens using ProtoScript II (New England Biolabs, Ipswich, MA). The Ion AmpliSeq SARS-CoV-2 Panel (Thermo-Fisher, Waltham, MA) was used to amplify 237 viral specific targets encompassing the complete viral genome. Libraries were sequenced and analyzed as we have previously described (9). For phylogenetic inference (i.e. to determine the hierarchy of case relationships), sequences were integrated with associated metadata and aligned on a local implementation of NextStrain (10) using augur and displayed via a web browser using auspice. Cases sequenced in this study were analyzed against a background collection of 1,120 SARS-CoV-2 genomes sequenced at Gundersen Health System between March and December 2020.

### Ethical approval

Specimens were analyzed in this study under a protocol approved by the Gundersen Health System Institutional Review Board (#2-20-03-008; PI: Kenny) to perform next-generation sequencing on remnant specimens after completion of diagnostic testing.

## RESULTS

To protect patient privacy, we will not disclose precise diagnosis dates but instead provide a numbered timeline centered on the date of the first diagnosis in the apparent cluster. The first patient case of COVID-19 in this investigation was diagnosed on a date between Nov 1-15, 2020, which we designate “Day 0”. One additional patient was diagnosed on day 1, two more followed on day 3 and a staff member was diagnosed on day 10. Hemodialysis patients typically utilize the facility once every two days, and all four COVID-19-positive patients shared the same alternate day schedule. These details and the treatment location for each individual are summarized on the facility map (Figure 1).

**Figure 1.**
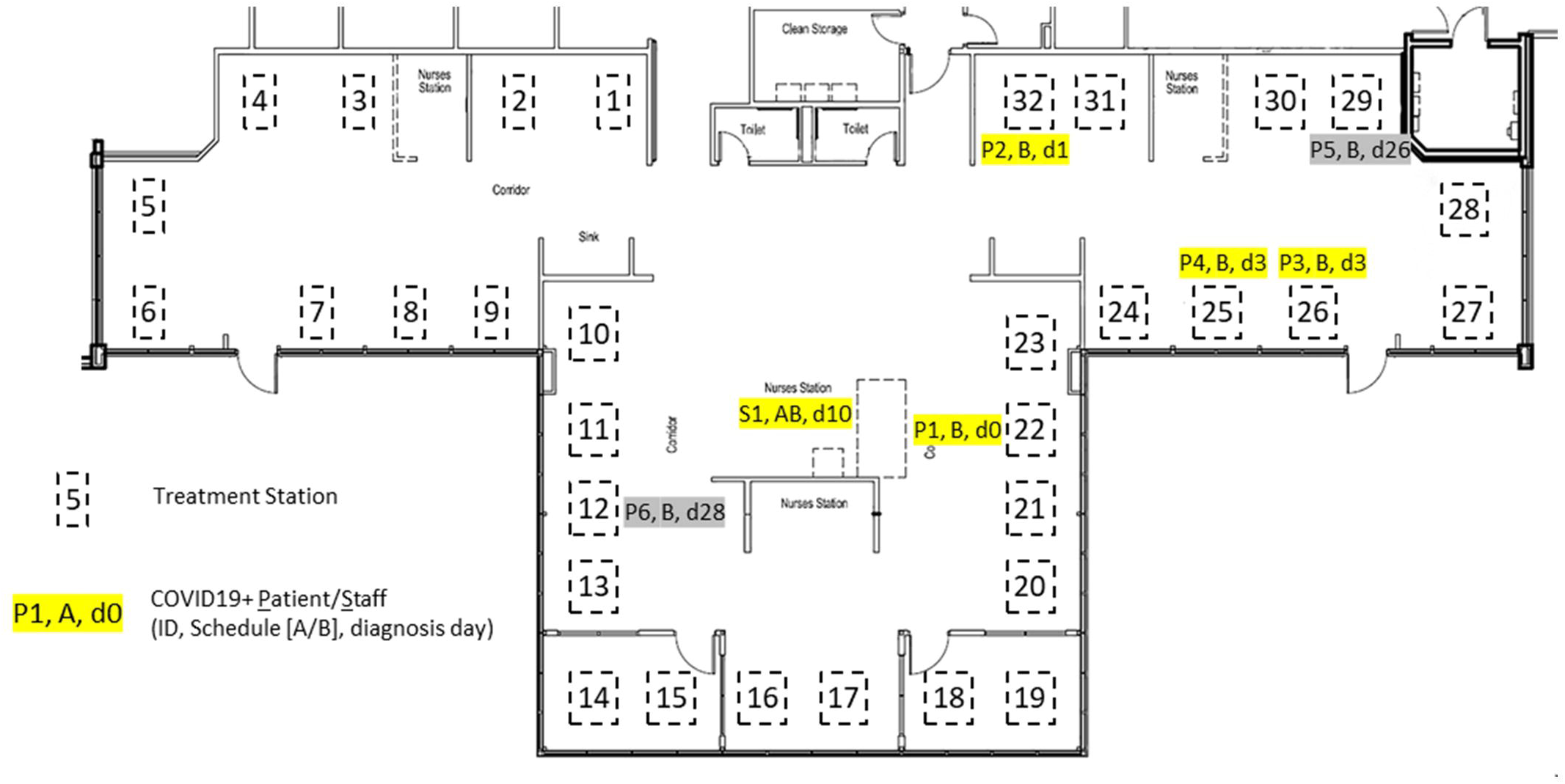
Schematic representation of the ESRD facility showing the location of treatment stations used by COVID-19-positive individuals. Cases are identified by an ID (P = Patient, S = Staff), which of two non-overlapping dialysis schedules was utilized (A or B) and the date of diagnosis relative to the diagnosis date of the initial patient of this cluster investigation. The five cases comprising the current cluster investigation are highlighted in yellow. Two cases that were detected subsequent to the current investigation are shown in gray.

The facility administrator and manager contacted local public health officials after the second, third and fifth cases. After the fifth case (day 10), health officials requested facility-wide testing of all patients and staff. This was performed on days 12 and 13 using the BinaxNOW antigen test. In the facility-wide testing 47 of 47 employees and 107 of 107 patients tested were negative. One patient refused testing and two patients were not present on either testing day.

The COVID-19 sequencing team was notified of the potential cluster on day 10. Of the five positive cases, four residual specimens were available for sequencing. Sequencing was completed on day 14. Genomes from each investigated specimen were compared to each other, and to a total of 1,120 genomes sequenced by the team from this region between March and December 2020. Each of the four samples analyzed was clearly genetically distinct from the others (Figure 2), unambiguously demonstrating that within-facility spread did not give rise to this apparent five case cluster.

**Figure 2.**
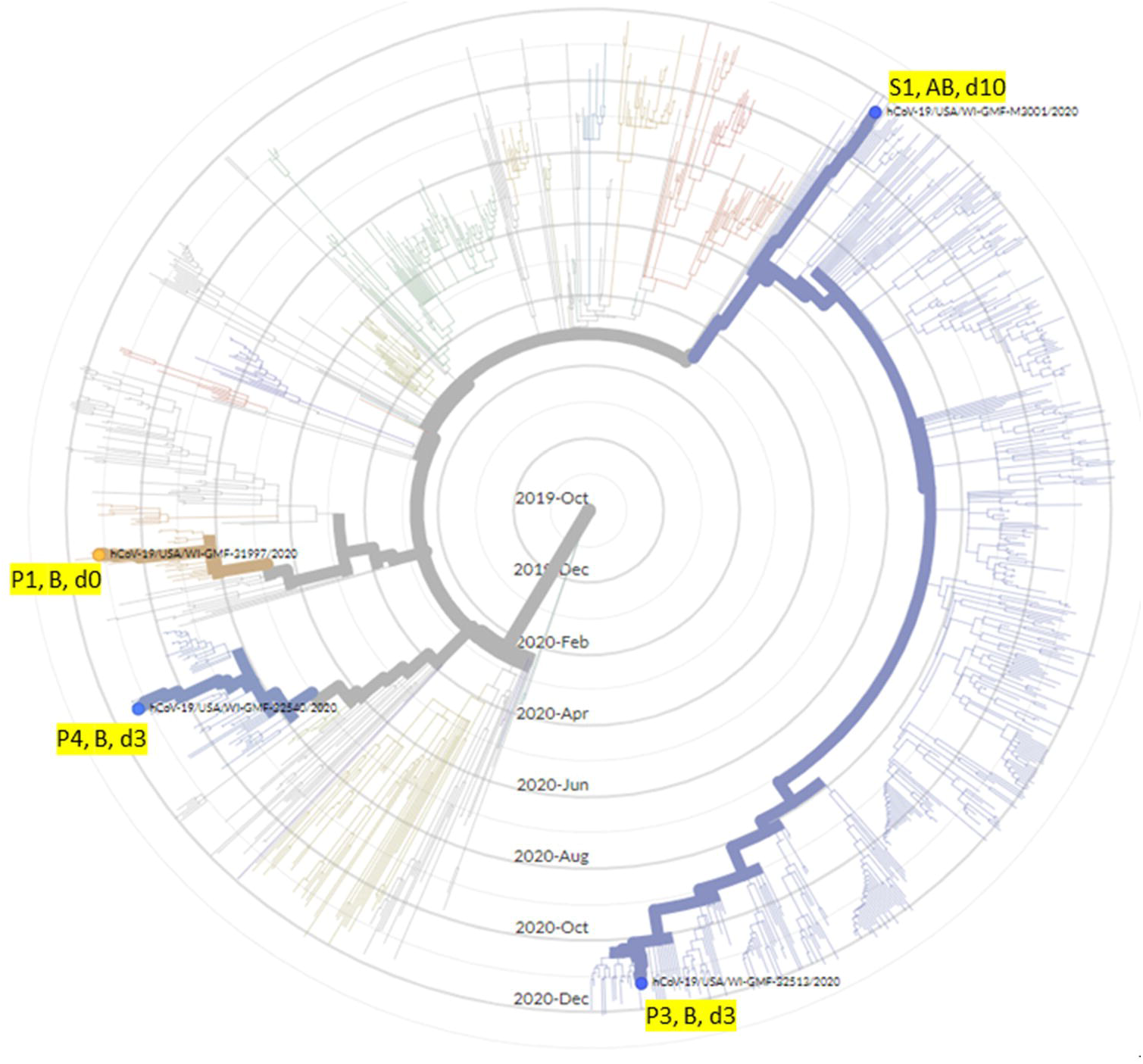
Four genetically distinct viral genomes sequenced from patient and staff in this ESRD cluster investigation. Radial phylogenetic tree representing 1,120 SARS-CoV-2 genomes sequenced at the Gundersen Health System between March – December 2020, with cases relevant to this cluster investigation highlighted. The tip of each branch represents a case and more genetically similar genomes cluster together. Specimens are identified using the code described in Figure 1.

Antigen testing (with confirmation by isothermal amplification for any positive cases) was chosen for the facility-wide screening for pragmatic reasons, most significantly (1) same-day result, preventing the need for patients or staff to isolate while results were pending, (2) the anterior nares swab was more acceptable to patients than the nasopharyngeal swab used for RT-PCR testing, (3) testing capacity constraints (154 additional RT-PCR tests in a single day was close to half of the daily throughput of the hospital’s laboratory) and (4) cost (an allowance of test kits provided at no-cost by the state of Wisconsin was used in this instance, obviating any delays due to potential disagreements on how screening costs should be assigned). Nevertheless, the false negative rate associated with antigen-directed testing among asymptomatic individuals (11) was concerning so we carefully monitored the ESRD facility in the following weeks to identify cases or spread that may have escaped surveillance using this particular assay. In the 14 days following the facility-wide testing, no additional staff cases were identified. Two patient additional cases were identified on days 26 and 28. Residual specimens were available for sequencing from these two cases, and analysis confirmed that they were distinct from each other and from the other cases described in this study.

## DISCUSSION

In this study, we demonstrate the contribution that rapid turnaround SARS-CoV-2 genome sequencing can make to infection cluster investigation. In this ESRD facility, five cases occurred within an eleven-day period, exceeding Wisconsin’s threshold for conducting a facility-wide investigation. The shared dialysis schedule and the proximity of the treatment stations for several of the affected individuals gave rise for additional concern about within-facility spread. While antigen testing of all patients and staff subsequently showed that SARS-CoV-2 infection was limited to only those individuals comprising the putative outbreak, the genomic analysis of the four available specimens conclusively demonstrated that these viruses each possessed distinct genomic lineages, and therefore could not have originated from a spread of a single viral substrain occurring within the limited window this cohort spent in the dialysis facility, as would be expected in a common source outbreak. Showing that SARS-CoV-2 spread had not occurred between the sequenced individuals in the facility refuted this presumptive cluster and provided staff and patients with reassurance that existing infection control procedures were working well.

We cannot exclude the possibility that the sample that was unavailable for sequencing (P2) might have matched one of the sequenced specimens. However, in the context of the otherwise negative facility-wide screening, the physical separation between P2’s treatment station and the nearest station used by a COVID19-positive individual (P4, Figure 1), the universal masking practiced in the facility, and P2’s other exposure risk factors (P2 lives in a congregate setting and requires care for complex medical needs), we considered it more likely than not that P2’s COVID-19 infection was unconnected to the other individuals.

When the first case of this investigation was identified (day 0), the county in which the facility is located reported a 7-day rolling average test positivity of 27% among symptomatic patients, with a known active case burden (cases identified within the previous 14 days) of 101 active cases per 10,000 residents. By day 11 when the fifth case was identified, the county’s level 7-day rolling average test positivity remained little changed at 29%, while the known active case burden had increased to 154 cases per 10,000 residents. In the presence of such widespread community activity, ESRD patients and staff may commonly acquire infections outside of the dialysis facility. While this places other patients at risk if institutional infection control procedures are weak, it may also lead to considerable over-burdening of institutional and public health resources investigating apparent clusters of cases that lack a common infection source.

The facility had implemented progressively more stringent face masking protocols beginning in April, and all patients and staff were masked at all times since July. During this investigation, it was determined that one of the positive patients who, had passed through screening multiple times, reported having had a “cough for a few weeks” when tested. This led to re-education of staff on the importance of diligent use of the screening tools at the entrance to the facility. Despite this screening failure, the absence of detected COVID-19 transmission from this individual in the facility over multiple visits underlines the value of educating patients and staff about mask use and enforcing these rules.

Nationwide, outbreaks in ESRD facilities have resulted in adverse impact to patients (morbidity and mortality among infected individuals, as well as disruption in dialysis schedules/locations for others). Public health recommendations include additional surveillance testing at weekly intervals for up to 28 days until there is a 7-day period of no positive cases. Using sequence data allowed us to demonstrate that this collection of cases was not a cluster of linked infections and therefore, the facility avoided the need for further rounds of surveillance screening of staff and patients. Although genetic analysis subsequently confirmed that these cases were not linked (three days after the decision to implement facility wide testing was taken), a more rapid demonstration that these cases were truly unlinked might have prevented the need for such extensive testing. Given the community case burden at the time of this study, it is likely that weekly surveillance testing may have continued to identify occasional sporadic cases, creating the false impression of a possible ongoing within-facility outbreak.

Though the risks of COVID-19 to ESRD patients are considerable, it is important to account also for the considerable resources involved in monitoring dialysis facilities during putative outbreaks. In our case, the rationale for performing facility-wide screening was clear and concordant with public health guidance. We posit that quicker access to sequencing data, and rapidly demonstrating the lack of a credible genetic and epidemiologic link in the cases in question, may spare the expenses of subsequent rounds of screening in instances similar to our own. Estimating using Medicare reimbursement rates, a single round of screening in a facility of this size would cost $7,700 (antigen testing) or $15,400 (RT-PCR). Conversely, sequencing costs per specimen (five each) amounted to approximately $200 dollars. In this proportion, the speed, cost, and surety provided by sequencing are compelling features that support its more regular inclusion in outbreak investigations as a way to conserve healthcare resources. Moreover, because resource shortages (e.g. PPE, testing capacity) have been an ongoing hallmark of the COVID-19 pandemic in the US, the public health response will likely benefit from redirection of such resources toward higher yield activities.

An adequate, trained and willing workforce as well as robust infection control training and procedures are recognized as key elements in institutional resilience against infectious disease outbreaks (12). Studies of healthcare workers who delivered care during the first SARS pandemic demonstrated long term adverse impacts (13), an experience that will likely be recapitulated at much larger scale in the current SARS-CoV-2 pandemic. Accordingly, it is important for staff to know that not every “outbreak” represents a collective failure to control disease spread. By demonstrating that this set of cases was not linked and did not lead to intra-facility spread, the genetic data strongly underlined the value of the infection control procedures that were practiced by both staff and patients. They confirm that the ESRD facility was a safe place in which to work and to receive care. Conversely, if the data had indicated some evidence of within-facility spread, the more granular nature of the genetic data may have led to the provision of targeted interventions to mitigate specific risk factors that would have been more challenging to identify from simply a numerical cluster of cases.

In conclusion, the exclusion of a true outbreak in our dialysis facility by way of robust genetic sequencing data validates the integrity of refined infection control practices in these critically important facilities, enabled provision of uninterrupted safe care to vulnerable patients in the midst of accelerating community spread, and highlighted the value of an interconnected network of nimble players in infection control, nursing, public health, and scientific laboratories. We anticipate that the benefits of this collaboration will serve as a model for the increasing use of rapid genomic sequencing data to shape institutional as well as public health responses in future outbreak scenarios in facilities of all sizes. As technology and expertise permit, we anticipate that the tools to quickly differentiate true outbreaks from pseudo-outbreaks will disseminate further into the healthcare landscape, and provide tangible benefits in other congregate settings.

## Data Availability

Sequence data are deposited in GISAID.

https://www.gisaid.org/

## ACKNOWLEDGEMENTS

This work was supported by the Gundersen Medical Foundation. PAK holds the Dr. Jon & Betty Kabara Endowed Chair in Precision Oncology.

